# Artificial Intelligence Predicts Health-Related Quality of Life for Adolescent Idiopathic Scoliosis

**DOI:** 10.1101/2025.11.16.25340349

**Authors:** Dusan Kovacevic, Aazad Abbas, Gurjovan Sahi, Johnathan R. Lex, Amer F. Samdani, Suken A. Shah, David Clements, Peter O. Newton, Michael P. Kelly, Jay Toor, Firoz Miyanji

## Abstract

**Purpose:** Adolescent idiopathic scoliosis (AIS) has a large impact on health-related quality of life (HRQoL) including poor psychosocial functioning, body image, and psychological distress. Surgical management for AIS is common; however, there is limited consensus on preoperative and intraoperative strategies to optimize HRQoL outcomes. Accurate prediction of postoperative outcomes can help guide operative planning and lead to improved HRQoL. This study aimed to generate machine learning models (MLMs) using preoperative and intraoperative variables to predict the difference in HRQoL outcomes from preoperative assessment to two years following AIS surgery.

**Methods:** A prospective, longitudinal, multicenter database was queried for AIS patients of Lenke 1 or 5 classification with two-year follow-up. MLMs were generated using preoperative and intraoperative variables to predict the difference in Scoliosis Research Society-22 (ΔSRS-22) questionnaire scores from preoperative assessment to two-year follow-up. MLMs were compared to a model that estimates the mean score by evaluating the mean squared error (MSE) and the fraction of times the prediction was within a predesignated value of the actual score (i.e., buffer accuracy).

**Results:** A total of 1,477 patients (84.6% female, 75.0% White) were included. The lowest MSE for each ΔSRS-22 outcome ranged from 0.18–0.48, while the highest 0.25-buffer, 0.5-buffer, 0.75-buffer, and 1-buffer accuracies for each ΔSRS-22 outcome ranged from 34.8%–53.4%, 56.8%–83.1%, 75.0%–94.6%, and 87.2%–97.3%, respectively. These MSEs and buffer accuracies outperformed mean estimates.

**Conclusion:** MLMs built using preoperative and intraoperative variables enabled prediction of the difference in HRQoL outcomes from preoperative assessment to two years following AIS surgery. Findings provide key insights into the feasibility of implementing MLMs to guide operative planning and counsel patients on expected outcomes of surgical management. Future work should implement additional surgeon, institution, and patient factors as model predictors to increase predictive accuracy of HRQoL outcomes and ultimately improve individualized patient care through data-driven surgical planning.

## 1. Introduction

Adolescent idiopathic scoliosis (AIS) is a structural deformity consisting of a lateral and rotated curvature of the spine that develops in adolescence without an obvious cause [1–3]. AIS has a large impact on health-related quality of life (HRQoL) including poorer psychosocial functioning, body image, and psychological distress [4–9].

Despite the well-established benefits of surgical intervention, determining a surgical treatment plan for AIS can be challenging given limited consensus on optimal operative strategies [10–15]. Previous studies documented large variability in surgical planning including fusion levels, type and density of implants used, degree of correction, use of biologics, and use of intraoperative traction between surgeons [16–23]. Due to the complexities and lack of clear guidelines, surgeons have relied on their training, experience, and clinical judgement to guide operative planning.

Considering the large impact of AIS on HRQoL, it is important to implement preoperative and intraoperative strategies that optimize HRQoL outcomes following surgery. The accurate prediction of postoperative HRQoL outcomes may help support surgeons with operative planning to maximize these outcomes [24]. Previous studies have predicted postoperative outcomes such as pain and satisfaction; however, these studies tend to be limited by smaller sample sizes and the inclusion of fewer predictors, which may affect generalizability of findings [25–27].

One potential tool to address these shortcomings and improve prediction of treatment outcomes is to utilize machine learning models (MLMs) built on large existing databases, which can incorporate diverse preoperative and intraoperative factors [28]. Generating comprehensive and applicable MLMs that accurately predict postoperative HRQoL outcomes may guide surgeons in counselling patients on expected outcomes of surgical management, inform surgical planning, and ultimately allow for optimal patient outcomes through data-driven surgical care.

To date, there is limited research implementing MLMs to predict HRQoL outcomes in patients undergoing surgical treatment of AIS. As such, this study aimed to generate MLMs using preoperative and intraoperative variables to predict the difference in HRQoL outcomes from preoperative assessment to two years following AIS surgery. A secondary objective was to identify key predictors of the top-performing MLMs.

## 2. Materials and Methods

### 2.1 Database and Study Population

This study was a retrospective analysis of prospectively collected data from the Harms Study Group founded by the Setting Scoliosis Straight Foundation [29]. The Harms Study Group prospectively collects data across paediatric and adolescent patients undergoing surgical treatment for spinal deformity across North America. Ethics approval was obtained from each institution and all patients consented for enrollment in the registry.

The database was queried for patients undergoing surgical correction of AIS with two-year follow-up. Lenke 1 or 5 curve types were included since these curves only have one structural region and therefore may be most suitable for predictive modeling. All lumbar spine modifiers and thoracic sagittal profiles were included. Patients missing HRQoL scores at two-year follow-up or missing more than 50% of data were excluded. Continuous variables were summarized with mean and standard deviation (SD), while categorical variables were summarized with total counts (N) and percentages (%).

### 2.2 Predictor and Outcome Variables

Preoperative and intraoperative variables were included as model predictors and are listed in Table 1. HRQoL was assessed preoperatively and two years postoperatively using the Scoliosis Research Society-22 (SRS-22) questionnaire [30]. The questionnaire consists of 22 questions covering five domains: pain, general function, self-image, mental health, and satisfaction. Total and domain scores range from 1–5, with higher scores indicating better patient outcomes. Model outcomes were characterized as the difference in SRS-22 total and individual domain scores from preoperative assessment to two-year follow-up (i.e., ΔSRS-22).

**Table 1.** Preoperative and intraoperative variables used to construct the machine learning models. Variable type Variable names (unit).

| Variable type | Variable names (unit) |
| --- | --- |
| Preoperative |  |
| Continuous | Age at AIS diagnosis (years), preoperative SRS-22 total, preoperative SRS-22 general function, preoperative SRS-22 pain, preoperative SRS-22 self-image, preoperative SRS-22 mental health, preoperative SRS-22 satisfaction, VAS pain, XR PA lumbar bend (°), XR lateral pelvic incidence (°), XR PA coronal C7 CVSL (°), XR lateral C7 sacrum (°), XR lateral lordosis T12 top sacrum (°), XR lateral pelvic tilt (°), XR PA Risser (°) |
| Categorical | Sex, race, smoking status, lumbar spine modifier, thoracic sagittal profile |
| Intraoperative |  |
| Continuous | Number of ribs removed, left rod size, right rod size, KSI of left rod, KSI of right rod |
| Categorical | Both ends locked before direct vertebral rotation, derotation with one rod in, derotation if one rod, derotation with two rods in, derotation if two rods, concave rod completely locked down before second rod put in, derotation maneuver performed, left/right rod material, left/right rod supplier, left/right system model, posterior based discectomy, concave rib osteotomy, ponte osteotomy, primary release lower level, primary release upper level, wide posterior release, posterior crosslink T2–L4, posterior fixation left T2–L3, posterior fixation right T2–L3, in-situ bending, rod tracing performed if no in-situ bending |

AIS: adolescent idiopathic scoliosis; SRS-22: Scoliosis Research Society-22; VAS: visual analogue scale; XR: x-ray; PA: posteroanterior; CVSL: central sacral vertical line.

### 2.3 Data Processing

The dataset was randomly split into training (80%) and testing sets (20%). Preoperative and intraoperative variables missing in more than 50% of patients were removed, with remaining variables included as model predictors. Missing continuous variables were imputed using an iterative imputer and missing categorical variables were imputed using the most common category [31]. Variables were appropriately scaled and adjusted to account for outliers. The correlation between each pair of variables was determined using Pearson’s correlation coefficient. Additional details on variable preprocessing methods are provided in Abbas et al [32].

### 2.4 MLM Construction

MLMs were generated using preoperative and intraoperative factors to predict ΔSRS-22 scores. Separate models were created for each of the six outcomes. MLMs used were linear regression [33], stochastic gradient descent (SGD) regression [34], K-nearest neighbours (KNN) [35], random forest [36], AdaBoost [37], and neural network [38]. To compare MLMs to a baseline model, a mean regressor that uses the mean of the training data as the outcome prediction for the test data was constructed. Models were trained and hyperparameters tuned to minimize mean squared error (MSE) using five-fold cross validation. The model with the smallest MSE on the testing dataset was selected as the top-performing MLM. If multiple models had equal MSEs, the model with the highest buffer accuracy was selected.

### 2.5 Outcome Metrics

MSE and buffer accuracy were used as outcome metrics to evaluate model quality [32].

Buffer accuracy was defined as the fraction of times the predicted outcome was within a predesignated value of the actual score. For instance, if the ΔSRS-22 score is 2, then a positive prediction occurs when the predicted difference is within +/- 0.25 of 2 (the actual score) for a 0.25-buffer. A higher buffer accuracy represents increased precision of the outcomes predicted by a model. The four predesignated buffers used were 0.25, 0.5, 0.75, and 1 to assess the precision of model estimates across different value ranges.

### 2.6 Identification of Predictors

The most important predictors for the top-performing MLMs were identified for all six HRQoL outcomes. Predictor importance was normalized with respect to the most important predictor in each model and presented in percentages as described by Abbas et al [32].

## 3. Results

A total of 1,477 patients (84.6% female, 75.0% White) were included in this study (training = 1,182, testing = 295). The average age at AIS diagnosis was 12.5 (SD 2.2) years. For the lumbar spine modifier, 692 (46.9%) patients were labelled as A, 245 (16.6%) as B, and 540 (36.6%) as C. For the thoracic sagittal profile, 234 (15.8%) patients were labelled as hypo, 1102 (74.6%) as normal, and 120 (8.1%) as hyper. Distributions of preoperative and intraoperative predictors used to construct the models are summarized in Appendices 1 and 2, respectively.

### 3.1 Model Results

Training and testing results for the models predicting HRQoL outcomes are shown in Tables 2–7.

**Table 2.** Training and testing results for the machine learning models predicting differences in SRS-22 total scores.

| Model Name | MSE | 0.25-Buffer Accuracy (%) | 0.5-Buffer Accuracy (%) | 0.75-Buffer Accuracy (%) | 1-Buffer Accuracy (%) |
| --- | --- | --- | --- | --- | --- |
| Training |  |  |  |  |  |
| Mean Estimates | 0.25 | 41.0 | 73.8 | 87.7 | 94.9 |
| Linear Regression | 0.14 | 53.6 | 86.3 | 95.9 | 98.1 |
| SGD | 0.25 | 41.0 | 73.8 | 87.7 | 94.9 |
| KNN | 0.16 | 53.2 | 82.6 | 93.8 | 98.0 |
| Random Forest | 0.06 | 79.9 | 95.5 | 98.5 | 99.2 |
| **AdaBoost** | 0.00 | 100.0 | 100.0 | 100.0 | 100.0 |
| Neural Network | 0.12 | 60.3 | 88.4 | 96.4 | 98.6 |
| Testing |  |  |  |  |  |
| Mean Estimates | 0.26 | 47.3 | 76.4 | 88.9 | 94.3 |
| Linear Regression | 0.19 | 49.0 | 76.4 | 91.6 | 97.0 |
| SGD | 0.26 | 47.3 | 76.4 | 88.9 | 94.3 |
| KNN | 0.25 | 44.6 | 74.3 | 88.2 | 96.0 |
| **Random Forest** | 0.18 | 48.3 | 83.1 | 94.6 | 97.3 |
| AdaBoost | 0.19 | 44.9 | 81.4 | 93.6 | 96.0 |
| **Neural Network** | 0.18 | 49.7 | 79.4 | 93.2 | 97.3 |

Model with the lowest MSE is bolded. SRS-22: Scoliosis Research Society-22; SGD: stochastic gradient descent; KNN: K-nearest neighbour; MSE: mean squared error.

**Table 3.** Training and testing results for the machine learning models predicting differences in SRS-22 general function scores.

| Model Name | MSE | 0.25-Buffer Accuracy (%) | 0.5-Buffer Accuracy (%) | 0.75-Buffer Accuracy (%) | 1-Buffer Accuracy (%) |
| --- | --- | --- | --- | --- | --- |
| Training |  |  |  |  |  |
| Mean Estimates | 0.37 | 37.9 | 63.8 | 83.1 | 89.8 |
| Linear Regression | 0.14 | 52.3 | 85.9 | 95.4 | 98.3 |
| SGD | 0.37 | 37.9 | 63.8 | 83.1 | 89.8 |
| KNN | 0.21 | 49.1 | 76.4 | 89.7 | 95.5 |
| Random Forest | 0.09 | 71.4 | 92.6 | 96.9 | 98.6 |
| AdaBoost | 0.02 | 91.0 | 100.0 | 100.0 | 100.0 |
| **Neural Network** | 0.00 | 99.5 | 100.0 | 100.0 | 100.0 |
| Testing |  |  |  |  |  |
| Mean Estimates | 0.38 | 40.5 | 68.2 | 82.8 | 89.5 |
| Linear Regression | 0.22 | 49.3 | 75.7 | 91.2 | 94.6 |
| SGD | 0.38 | 40.5 | 68.2 | 82.8 | 89.5 |
| KNN | 0.38 | 40.2 | 66.2 | 81.1 | 90.5 |
| **Random Forest** | 0.20 | 49.0 | 79.7 | 92.2 | 95.3 |
| **AdaBoost** | 0.20 | 53.4 | 81.4 | 92.2 | 96.3 |
| Neural Network | 0.24 | 48.3 | 74.7 | 88.9 | 94.6 |

Model with the lowest MSE is bolded. SRS-22: Scoliosis Research Society-22; SGD: stochastic gradient descent; KNN: K-nearest neighbour; MSE: mean squared error.

**Table 4.** Training and testing results for the machine learning models predicting differences in SRS-22 mental health scores.

| Model Name | MSE | 0.25-Buffer Accuracy (%) | 0.5-Buffer Accuracy (%) | 0.75-Buffer Accuracy (%) | 1-Buffer Accuracy (%) |
| --- | --- | --- | --- | --- | --- |
| Training |  |  |  |  |  |
| Mean Estimates | 0.56 | 36.4 | 56.7 | 71.1 | 84.3 |
| Linear Regression | 0.29 | 38.0 | 66.7 | 84.3 | 93.3 |
| SGD | 0.56 | 36.4 | 56.7 | 71.1 | 84.3 |
| KNN | 0.38 | 35.7 | 64.4 | 81.9 | 89.8 |
| Random Forest | 0.10 | 64.0 | 90.5 | 96.8 | 98.7 |
| **AdaBoost** | 0.00 | 100.0 | 100.0 | 100.0 | 100.0 |
| Neural Network | 0.06 | 78.6 | 94.4 | 98.7 | 99.6 |
| Testing |  |  |  |  |  |
| Mean Estimates | 0.61 | 33.8 | 55.1 | 70.3 | 82.8 |
| Linear Regression | 0.55 | 27.7 | 55.1 | 72.0 | 83.5 |
| SGD | 0.61 | 33.8 | 55.1 | 70.3 | 82.8 |
| KNN | 0.60 | 29.4 | 53.7 | 72.0 | 81.8 |
| **Random Forest** | 0.48 | 29.4 | 56.8 | 75.0 | 86.8 |
| **AdaBoost** | 0.48 | 34.8 | 56.4 | 73.7 | 87.2 |
| Neural Network | 0.54 | 26.4 | 49.0 | 71.3 | 84.8 |

Model with the lowest MSE is bolded. SRS-22: Scoliosis Research Society-22; SGD: stochastic gradient descent; KNN: K-nearest neighbour; MSE: mean squared error.

**Table 5.** Training and testing results for the machine learning models predicting differences in SRS-22 pain scores.

| Model Name | MSE | 0.25-Buffer Accuracy (%) | 0.5-Buffer Accuracy (%) | 0.75-Buffer Accuracy (%) | 1-Buffer Accuracy (%) |
| --- | --- | --- | --- | --- | --- |
| Training |  |  |  |  |  |
| Mean Estimates | 0.60 | 22.0 | 54.3 | 73.8 | 82.6 |
| Linear Regression | 0.28 | 40.0 | 70.2 | 86.6 | 93.7 |
| SGD | 0.60 | 22.0 | 54.3 | 73.8 | 82.6 |
| KNN | 0.38 | 37.4 | 62.9 | 79.9 | 89.5 |
| Random Forest | 0.15 | 54.6 | 84.7 | 93.8 | 97.3 |
| **AdaBoost** | 0.00 | 100.0 | 100.0 | 100.0 | 100.0 |
| Neural Network | 0.01 | 98.3 | 99.8 | 99.9 | 100.0 |
| Testing |  |  |  |  |  |
| Mean Estimates | 0.55 | 19.6 | 56.1 | 74.0 | 83.1 |
| Linear Regression | 0.41 | 33.5 | 62.5 | 78.4 | 89.2 |
| SGD | 0.55 | 19.6 | 56.1 | 74.0 | 83.1 |
| KNN | 0.55 | 31.4 | 51.4 | 68.2 | 83.1 |
| **Random Forest** | 0.37 | 39.2 | 60.5 | 82.1 | 91.6 |
| AdaBoost | 0.38 | 41.6 | 63.9 | 78.7 | 91.2 |
| Neural Network | 0.51 | 31.1 | 56.1 | 74.3 | 86.5 |

Model with the lowest MSE is bolded. SRS-22: Scoliosis Research Society-22; SGD: stochastic gradient descent; KNN: K-nearest neighbour; MSE: mean squared error.

**Table 6.** Training and testing results for the machine learning models predicting differences in SRS-22 satisfaction scores.

| Model Name | MSE | 0.25-Buffer Accuracy (%) | 0.5-Buffer Accuracy (%) | 0.75-Buffer Accuracy (%) | 1-Buffer Accuracy (%) |
| --- | --- | --- | --- | --- | --- |
| Training |  |  |  |  |  |
| Mean Estimates | 1.13 | 18.4 | 35.4 | 49.0 | 68.5 |
| Linear Regression | 0.35 | 36.5 | 68.3 | 86.3 | 92.9 |
| SGD | 1.13 | 18.4 | 35.4 | 49.0 | 68.5 |
| KNN | 0.75 | 21.9 | 44.5 | 62.7 | 76.8 |
| Random Forest | 0.10 | 69.9 | 92.7 | 96.7 | 98.5 |
| **AdaBoost** | 0.00 | 99.4 | 100.0 | 100.0 | 100.0 |
| Neural Network | 0.01 | 96.2 | 99.0 | 99.8 | 99.8 |
| Testing |  |  |  |  |  |
| Mean Estimates | 1.21 | 18.5 | 34.6 | 47.7 | 66.9 |
| Linear Regression | 0.57 | 30.8 | 53.9 | 74.2 | 86.9 |
| SGD | 1.21 | 18.5 | 34.6 | 47.7 | 66.9 |
| KNN | 1.08 | 20.8 | 36.2 | 53.9 | 71.2 |
| **Random Forest** | 0.48 | 32.7 | 61.9 | 79.2 | 90.0 |
| AdaBoost | 0.50 | 37.3 | 80.4 | 82.3 | 92.7 |
| Neural Network | 0.66 | 25.4 | 53.1 | 70.4 | 83.5 |

Model with the lowest MSE is bolded. SRS-22: Scoliosis Research Society-22; SGD: stochastic gradient descent; KNN: K-nearest neighbour; MSE: mean squared error.

**Table 7.** Training and testing results for the machine learning models predicting differences in SRS-22 self-image scores.

| Model Name | MSE | 0.25-Buffer Accuracy (%) | 0.5-Buffer Accuracy (%) | 0.75-Buffer Accuracy (%) | 1-Buffer Accuracy (%) |
| --- | --- | --- | --- | --- | --- |
| Training |  |  |  |  |  |
| Mean Estimates | 0.60 | 20.7 | 49.5 | 71.8 | 81.9 |
| Linear Regression | 0.24 | 40.2 | 74.1 | 89.8 | 95.7 |
| SGD | 0.60 | 20.7 | 49.5 | 71.8 | 81.9 |
| KNN | 0.38 | 31.8 | 61.1 | 79.2 | 90.4 |
| Random Forest | 0.15 | 59.4 | 89.3 | 95.6 | 97.6 |
| AdaBoost | 0.05 | 81.1 | 99.2 | 100.0 | 100.0 |
| **Neural Network** | 0.01 | 97.6 | 99.8 | 100.0 | 100.0 |
| Testing |  |  |  |  |  |
| Mean Estimates | 0.59 | 19.3 | 51.7 | 71.6 | 81.8 |
| Linear Regression | 0.35 | 34.5 | 63.5 | 83.1 | 91.2 |
| SGD | 0.59 | 19.3 | 51.7 | 71.6 | 81.8 |
| KNN | 0.57 | 29.1 | 49.3 | 66.9 | 82.8 |
| **Random Forest** | 0.32 | 34.5 | 65.2 | 84.8 | 93.6 |
| AdaBoost | 0.34 | 37.2 | 63.9 | 81.4 | 94.6 |
| Neural Network | 0.42 | 33.8 | 59.5 | 77.7 | 90.9 |

Model with the lowest MSE is bolded. SRS-22: Scoliosis Research Society-22; SGD: stochastic gradient descent; KNN: K-nearest neighbour; MSE: mean squared error.

#### 3.1.1 Model Training

The AdaBoost model had the best performance across all metrics for ΔSRS-22 total, mental health, pain, and satisfaction scores, while the neural network model had the best performance across all metrics for general function and self-image scores.

#### 3.1.2 Model Testing

For ΔSRS-22 total scores, the random forest and neural network models had the lowest MSE (0.18) and highest 1-buffer accuracy (97.3%). The random forest model had the highest 0.5-buffer (83.1%) and 0.75-buffer accuracies (94.6%), while the neural network model had the highest 0.25-buffer accuracy (49.7%). These buffer accuracies outperformed mean estimates by 2.4%–6.7%.

For ΔSRS-22 general function scores, the random forest and AdaBoost models had the lowest MSE (0.20) and highest 0.75-buffer accuracy (92.2%). The AdaBoost model had the highest 0.25-buffer (53.4%), 0.5-buffer (81.4%), and 1-buffer accuracies (96.3%). These buffer accuracies outperformed mean estimates by 6.8%–13.2%.

For ΔSRS-22 mental health scores, the random forest and AdaBoost models had the lowest MSE (0.48). The random forest model had the highest 0.5-buffer (56.8%) and 0.75-buffer accuracies (75.0%), while the AdaBoost model had the highest 0.25-buffer (34.8%) and 1-buffer accuracies (87.2%). These buffer accuracies outperformed mean estimates by 1.0%–4.7%.

For ΔSRS-22 pain scores, the random forest model had the best MSE (0.37), 0.75-buffer (82.1%), and 1-buffer accuracies (91.6%), while the AdaBoost model had the highest 0.25-buffer (41.6%) and 0.5-buffer accuracies (63.9%). These buffer accuracies outperformed mean estimates by 7.8%–22.0%.

For ΔSRS-22 satisfaction scores, the random forest model had the lowest MSE (0.48). The highest 0.25-buffer (37.3%), 0.5-buffer (80.4%), 0.75-buffer (82.3%), and 1-buffer accuracies (92.7%) were achieved by the AdaBoost model, which outperformed mean estimates by 18.9%–45.8%.

For ΔSRS-22 self-image scores, the random forest model had the best MSE (0.32), 0.5-buffer (65.2%), and 0.75-buffer accuracies (84.8%), while the AdaBoost model had the highest 0.25-buffer (37.2%) and 1-buffer accuracies (94.6%). These buffer accuracies outperformed mean estimates by 12.8%–17.9%.

Comparisons of the predicted versus actual differences in HRQoL outcomes for the top-performing model of each outcome are displayed in Figure 1.

**Figure 1.**
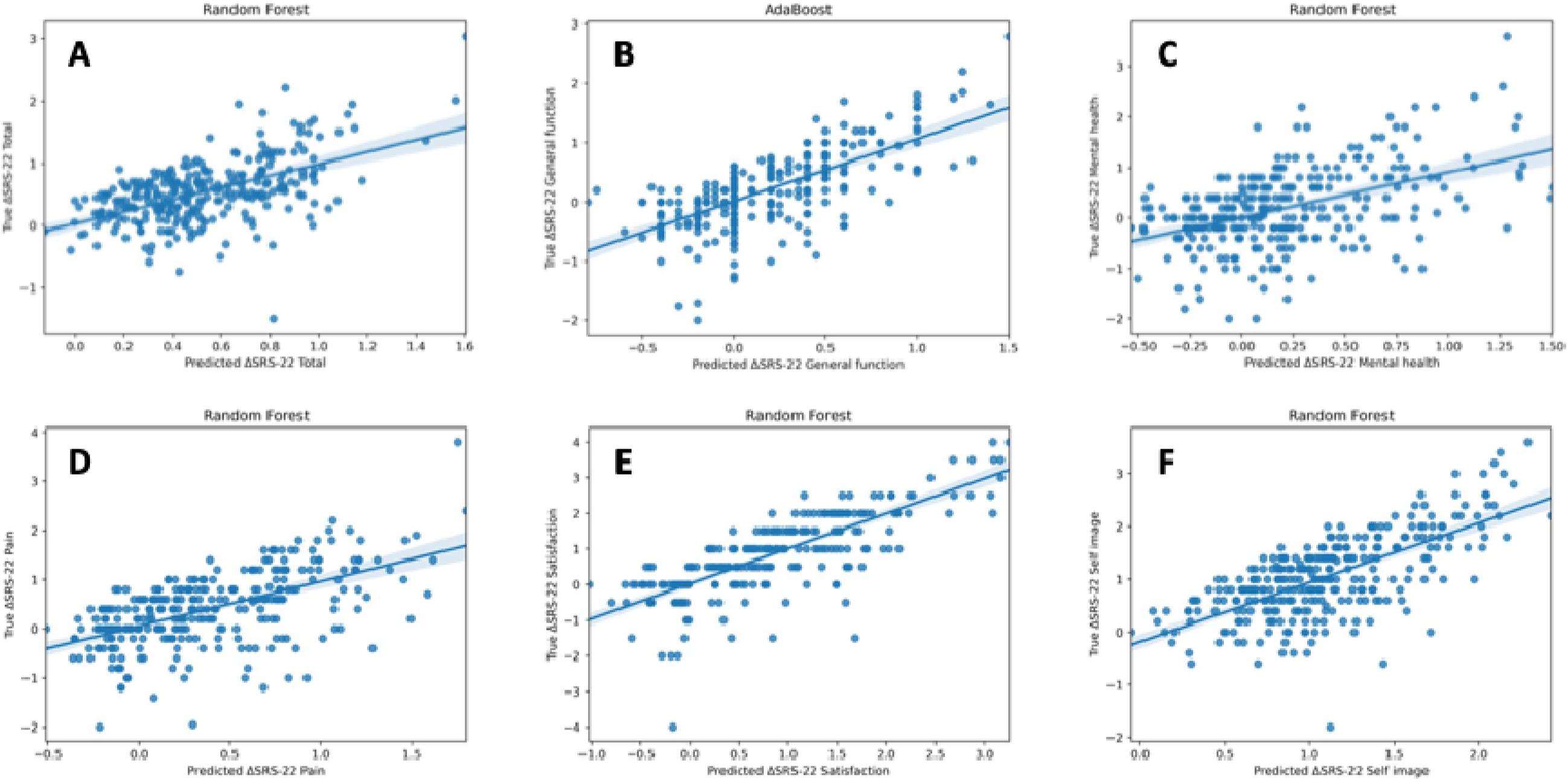
Predicted versus actual differences in Scoliosis Research Society-22 (ΔSRS-22) total and domain scores for the top-performing model of each outcome. A: Random forest model for ΔSRS-22 total; B: AdaBoost model for ΔSRS-22 general function; C: Random forest model for ΔSRS-22 mental health; D: Random forest model for ΔSRS-22 pain; E: Random forest model for ΔSRS-22 satisfaction; F: Random forest model for ΔSRS-22 self-image.

### 3.2 Predictor Importance

The top ten predictors of the top-performing MLM for each outcome are displayed in Table 8. The most important predictor across each outcome was the preoperative SRS-22 score of the respective domain (i.e., preoperative satisfaction score for the satisfaction outcome). Other important predictors that were common across outcomes included preoperative SRS-22 scores of different domains and preoperative radiographic variables such as lateral lordosis T12 top sacrum and posteroanterior lumbar bend.

**Table 8.** Top ten predictors of the top-performing models for each outcome.

| Total | General Function | Mental Health | Pain | Satisfaction | Self-Image |
| --- | --- | --- | --- | --- | --- |
| Random Forest | AdaBoost | Random Forest | Random Forest | Random Forest | Random Forest |
| SRS-22 total (100) | SRS-22 general function (100) | SRS-22 mental health (100) | SRS-22 pain (100) | SRS-22 satisfaction (100) | SRS-22 self-image (100) |
| XR lateral lordosis T12 top sacrum (11) | SRS-22 total (8) | XR PA coronal C7 CSVL (12) | VAS pain (10) | XR lateral lordosis T12 top sacrum (3) | SRS-22 mental health (8) |
| Age at AIS diagnosis (9) | SRS-22 pain (8) | SRS-22 total (12) | SRS-22 mental health (8) | SRS-22 total (3) | SRS-22 total (7) |
| SRS-22 pain (8) | XR lateral C7 sacrum (6) | SRS-22 general function (10) | Lumbar spine modifier (2) | VAS pain (3) | Number of ribs removed (7) |
| SRS-22 mental health (8) | SRS-22 satisfaction (6) | XR PA lumbar bend (10) | Race black (1) | SRS-22 pain (2) | SRS-22 general function (5) |
| SRS-22 general function (7) | SRS-22 mental health (6) | Age at AIS diagnosis (9) | No Ponte osteotomy (1) | SRS-22 general function (2) | SRS-22 pain (5) |
| SRS-22 satisfaction (7) | SRS-22 self-image (5) | SRS-22 satisfaction (7) | No posterior fixation left T6 (1) | SRS-22 mental health (2) | SRS-22 satisfaction (4) |
| KSI of right rod (5) | XR lateral pelvic tilt (4) | KSI of right rod (5) | Left rod size (1) | XR PA lumbar bend (2) |  |
| XR PA Risser (5) | KSI of right rod (4) | XR PA Risser (4) | Race white (1) | XR lateral C7 sacrum (2) |  |
| KSI of left rod (4) | Posterior crosslink T11 (3) | KSI of left rod (3) | Primary release upper level T6 (1) | XR lateral pelvic tilt (2) |  |

Predictor importance normalized to the most important predictor in each model and presented as percentage. All SRS-22 predictors represent the preoperative scores. For the self-image outcome, only seven predictors were relevant. SGD: stochastic gradient descent; SRS-22: Scoliosis Research Society-22; XR: x-ray; PA: posteroanterior; CSVL: central sacral vertical line; AIS: adolescent idiopathic scoliosis; VAS: visual analogue scale.

## 4. Discussion

The present study generated MLMs using preoperative and intraoperative variables to predict ΔSRS-22 outcomes in patients undergoing surgical treatment of AIS. This work has profound implications in improving patient outcomes as findings highlight the potential for implementing MLMs to guide preoperative counselling, enhance surgical planning, and deliver data-driven surgical care.

The top 0.25-buffer, 0.5-buffer, 0.75-buffer, and 1-buffer accuracies for each HRQoL outcome ranged from 34.8%–53.4%, 56.8%–83.1%, 75.0%–94.6%, and 87.2%–97.3%, respectively. The top-performing models all had higher buffer accuracies compared to mean estimates, with improvements in accuracy ranging from 1.0%–45.8%. When comparing the top models across outcomes, the highest buffer accuracies were achieved for ΔSRS-22 total and general function scores, while the greatest improvements in accuracy relative to mean estimates were achieved for ΔSRS-22 satisfaction scores. The lowest buffer accuracies and smallest improvements in accuracy compared to mean estimates were achieved for ΔSRS-22 mental health scores. These findings suggest that MLMs may be most beneficial in predicting satisfaction, while mental health may be more difficult to predict. This is consistent with previous work that demonstrated MLMs perform better in predicting postoperative function, pain, and satisfaction, compared to mental health and self-image among AIS patients [39].

A key finding of this study was that the most important predictor for the top-performing model across each outcome was the preoperative SRS-22 score of the respective domain. Other important predictors included preoperative SRS-22 scores of other domains, age at AIS diagnosis, and preoperative radiographic variables. Previous work has also found preoperative radiographic variables and preoperative SRS-22 scores to be important predictors for postoperative patient satisfaction [39]. These findings highlight the importance of the preoperative clinical and radiographic assessment in guiding management of AIS.

While various preoperative and intraoperative variables were used as predictors in the MLMs, many other factors may contribute to postoperative HRQoL outcomes. For instance, surgeon and hospital characteristics such as surgeon experience, surgical team, and hospital infrastructure influence surgical outcomes but were not captured in the models [40–44].

Similarly, patient-specific factors like self-efficacy, social support, and resilience can contribute to patients’ wellbeing following surgical treatment and thus affect postoperative SRS-22 scores [45, 46]. Implementing these factors in future MLMs may improve predictive accuracy of HRQoL outcomes.

The implementation of MLMs that accurately predict HRQoL outcomes in patients undergoing surgical treatment of AIS may assist surgeons in developing personalized surgical plans by identifying optimal operative strategies to maximize HRQoL for each patient. Support with operative planning is particularly beneficial given the complexities, lack of formal guidelines, and limited consensus associated with surgical treatment for AIS [17]. In addition, MLMs can help with counselling patients on expected outcomes of surgical management and guide their decision making [28]. Patients at risk for worse outcomes can also be identified and provided with additional support to improve postoperative care. For example, a patient predicted to experience greater postoperative pain can be provided with a rehabilitation plan targeting pain reduction following surgery.

This study has several limitations. While the SRS-22 questionnaire is commonly used to evaluate HRQoL in AIS, it is limited by significant ceiling effects, which may reduce predictive accuracy particularly for patients with higher preoperative scores [47, 48]. However, a normal distribution transformation was applied to allow for optimal machine learning by diminishing the impact of the lower ceiling with this measure. This study also examined postoperative HRQoL outcomes at a two-year follow-up. Predictive abilities of MLMs may differ beyond the two-year mark as HRQoL of postoperative AIS patients tends to remain relatively stable within the first five years and then steadily decrease after 10 years [49]. Finally, this study included patients with Lenke 1 or 5 curve types as they only have one structural region. Other Lenke classifications have at least two structural regions, which may reduce predictive accuracy.

Additional work is needed to determine whether MLMs can be applied to other curve classifications.

## 5. Conclusion

This study generated MLMs using preoperative and intraoperative variables to predict the difference in HRQoL outcomes from preoperative assessment to two years following AIS surgery. The most important predictors of the top-performing models across outcomes were preoperative subjective HRQoL scores. Findings provide key insights into the feasibility of implementing MLMs to guide operative planning and counsel patients on expected outcomes of surgical management. Future work should implement additional surgeon, institution, and patient factors as model predictors to increase predictive accuracy of HRQoL outcomes and ultimately improve individualized patient care through data-driven surgical planning.

## Data Availability

All data produced in the present study are available upon reasonable request to the authors.

## Acknowledgement

This study was supported in part by grants to the Setting Scoliosis Straight Foundation in support of Harms Study Group research from DePuy Synthes Spine, Atec, Stryker Spine, Medtronic, Globus-NuVasive, Highridge, Biedermann Motech, Orthopediatrics, the Food and Drug Administration and Pediatric Orthopedic Society of North America.

## Harms Study Group Investigators

Aaron Buckland, MD; Melbourne Orthopaedic Grp & Royal Childrens Hosp.

Ahmet Alanay, MD; Acibadem Maslak Hospital, Turkey

Amer Samdani, MD; Shriners Hospitals for Children—Philadelphia

Amit Jain, MD; Johns Hopkins Hospital

Baron Lonner, MD; Mount Sinai Hospital

Benjamin Roye, MD; Columbia University

Bob Cho, MD; Shriner’s Pasadena CA

Burt Yaszay, MD; Seattle Children’s Hospital

Caglar Yilgor, MD; Acibadem Maslak Hospital, Turkey

Dan Hoernschmeyer, MD; University of Missouri Health Care

Daniel Hedequist, MD; Boston Children’s Hospital

Daniel Sucato, MD; Texas Scottish Rite Hospital

David Clements, MD; Cooper Bone & Joint Institute New Jersey

Firoz Miyanji, MD; BC Children’s Hospital

Harry Shufflebarger, MD; Paley Orthopedic & Spine Institute

Jack Flynn, MD; Children’s Hospital of Philadelphia

Jean Marc Mac Thiong, MD; CHU Sainte-Justine

Josh Murphy, MD; Children’s Healthcare of Atlanta

Joshua Pahys, MD; Shriners Hospitals for Children—Philadelphia

Keith Bachmann, MD; University of Virginia

Kevin Neal, MD; Nemours Children’s Clinic, Jacksonville

Laurel Blakemore, MD; Pediatric Specialists of Virginia/Children’s National

Lawrence Haber, MD; Ochsner Health Center for Children New Orleans

Lawrence Lenke, MD; Columbia University

Mark Abel, MD; University of Virginia

Mark Erickson, MD; Children’s Hospital, Denver Colorado

Michael Glotzbecker, MD; Rainbow Children’s Hospital, Cleveland

Michael Kelly, MD; Rady Children’s Hospital

Michael Vitale, MD; Columbia University

Michelle Marks, PT, MA; Setting Scoliosis Straight Foundation

Munish Gupta, MD; Washington University

Nicholas Fletcher, MD; Emory University

Noelle Larson, MD; Mayo Clinic Rochester Minnesota

Patrick Cahill, MD; Children’s Hospital of Philadelphia

Paul Sponseller, MD; Johns Hopkins Hospital

Peter Gabos, MD: Nemours/Alfred I. duPont Hospital for Children

Peter Newton, MD; Rady Children’s Hospital

Peter Sturm, MD; Cincinnati Children’s Hospital

Randal Betz, MD; Institute for Spine & Scoliosis

Stefan Parent, MD: CHU Sainte-Justine

Stephen George, MD; Nicklaus Children’s Hospital

Steven Hwang, MD; Shriners Hospitals for Children—Philadelphia

Suken Shah, MD; Nemours/Alfred I. duPont Hospital for Children

Sumeet Garg, MD; Children’s Hospital, Denver Colorado

Tom Errico, MD; Nicklaus Children’s Hospital

Vidyadhar Upasani, MD; Rady Children’s Hospital

