## Appendix 1 for "Artificial Intelligence Predicts Health-Related Quality of Life for Adolescent Idiopathic Scoliosis"

**Appendix 1.** Distributions of preoperative predictors used to construct the machine learning models.

| Predictor | | | Mean (SD) or N (%) |
| --- | --- | --- | --- |
| Continuous | | |  |
|  | Age at AIS diagnosis (years) | | 12.5 (2.2) |
|  | Preoperative SRS-22 total | | 3.9 (0.5) |
|  | Preoperative SRS-22 general function | | 4.4 (0.6) |
|  | Preoperative SRS-22 pain | | 4.0 (0.8) |
|  | Preoperative SRS-22 self-image | | 3.3 (0.7) |
|  | Preoperative SRS-22 mental health | | 4.0 (0.7) |
|  | Preoperative SRS-22 satisfaction | | 3.7 (1.0) |
|  | VAS pain | | 1.7 (2.4) |
|  | XR PA lumbar bend (°) | | 12.1 (8.9) |
|  | XR lateral pelvic incidence (°) | | 52.4 (13.1) |
|  | XR PA coronal C7 CSVL (°) | | -0.3 (2.3) |
|  | XR lateral C7 sacrum (°) | | -0.6 (3.7) |
|  | XR lateral lordosis T12 top sacrum (°) | | -57.7 (15.2) |
|  | XR lateral pelvic tilt (°) | | 9.0 (8.1) |
|  | XR PA Risser (°) | | 3.2 (1.4) |
| Categorical | | |  |
|  | Sex | |  |
|  |  | Female | 1249 (84.6) |
|  |  | Male | 226 (15.3) |
|  | Race | |  |
|  |  | White | 1108 (75.0) |
|  |  | Hispanic | 182 (12.3) |
|  |  | Black | 164 (11.1) |
|  | Smoking status | |  |
|  |  | Non-smoker | 1477 (100.0) |
|  | Lumbar spine modifier | |  |
|  |  | A | 692 (46.9) |
|  |  | B | 245 (16.6) |
|  |  | C | 540 (36.6) |
|  | Thoracic sagittal profile | |  |
|  |  | Hypo | 234 (15.8) |
|  |  | Normal | 1102 (74.6) |
|  |  | Hyper | 120 (8.1) |

Continuous predictors are summarized with mean and standard deviation (SD) and categorical predictors with total counts (N) and percentages (%). AIS: adolescent idiopathic scoliosis; SRS-22: Scoliosis Research Society-22; VAS: visual analogue scale; XR: x-ray; PA: posteroanterior; CSVL: central sacral vertical line.
