## Appendix 2 for "Artificial Intelligence Predicts Health-Related Quality of Life for Adolescent Idiopathic Scoliosis"

**Appendix 2.** Distributions of intraoperative predictors used to construct the machine learning models.

| Predictor | | | Mean (SD) or N (%) |
| --- | --- | --- | --- |
| Continuous | | |  |
|  | Number of ribs removed | | 2.0 (2.2) |
|  | Left rod size | | 5.5 (0.2) |
|  | Right rod size | | 5.5 (0.2) |
|  | KSI of left rod | | 181.9 (21.4) |
|  | KSI of right rod | | 176.2 (25.6) |
| Categorical | | |  |
|  | Both ends locked before direct vertebral rotation | |  |
|  |  | Yes | 175 (11.8) |
|  |  | No | 521 (35.3) |
|  |  | Null | 61 (4.1) |
|  | Derotation with one rod in | |  |
|  |  | Yes | 418 (28.3) |
|  |  | No | 326 (22.1) |
|  | Derotation if one rod | |  |
|  |  | En bloc | 140 (9.5) |
|  |  | Segmental | 132 (8.9) |
|  |  | Both | 108 (7.3) |
|  | Derotation with two rods in | |  |
|  |  | Yes | 526 (35.6) |
|  |  | No | 196 (13.3) |
|  | Derotation if two rods | |  |
|  |  | En bloc | 49 (3.3) |
|  |  | Segmental | 377 (25.5) |
|  |  | Both | 60 (4.1) |
|  | Concave rod completely locked down before second rod put in | |  |
|  |  | Yes | 150 (10.2) |
|  |  | No | 544 (36.8) |
|  |  | Null | 56 (3.8) |
|  | Derotation maneuver performed | |  |
|  |  | Yes | 1353 (91.6) |
|  | Left rod material | |  |
|  |  | Stainless steel | 736 (48.8) |
|  |  | Cobalt-chromium | 545 (36.9) |
|  |  | Titanium | 182 (12.3) |
|  | Left rod supplier | |  |
|  |  | DePuy | 1466 (99.3) |
|  | Left system model | |  |
|  |  | Expedium | 1461 (98.9) |
|  | Right rod material | |  |
|  |  | Stainless steel | 734 (49.7) |
|  |  | Cobalt-chromium | 538 (36.4) |
|  |  | Titanium | 191 (12.9) |
|  | Right rod supplier | |  |
|  |  | DePuy | 1466 (99.3) |
|  | Right system model | |  |
|  |  | Expedium | 1461 (98.9) |
|  | Posterior based discectomy | |  |
|  |  | No | 781 (52.9) |
|  | Concave rib osteotomy | |  |
|  |  | No | 1402 (94.9) |
|  | Ponte osteotomy | |  |
|  |  | Yes | 810 (54.8) |
|  |  | No | 470 (31.8) |
|  | Primary release lower level | |  |
|  |  | T9 | 61 (4.1) |
|  |  | T10 | 302 (20.4) |
|  |  | T11 | 186 (12.6) |
|  |  | T12 | 139 (9.4) |
|  |  | L1 | 68 (4.6) |
|  |  | L2 | 74 (5.0) |
|  |  | L3 | 129 (8.7) |
|  | Primary release upper level | |  |
|  |  | T4 | 34 (2.3) |
|  |  | T5 | 89 (6.0) |
|  |  | T6 | 360 (24.4) |
|  |  | T7 | 214 (14.5) |
|  |  | T8 | 74 (5.0) |
|  |  | T10 | 47 (3.2) |
|  |  | T11 | 75 (5.1) |
|  |  | T12 | 67 (4.5) |
|  | Wide posterior release | |  |
|  |  | Yes | 227 (15.4) |
|  |  | No | 1060 (71.8) |
|  | Posterior crosslink T2 | |  |
|  |  | False | 1206 (81.7) |
|  | Posterior crosslink T3 | |  |
|  |  | False | 1268 (85.9) |
|  | Posterior crosslink T4 | |  |
|  |  | False | 1380 (93.4) |
|  | Posterior crosslink T5 | |  |
|  |  | False | 1437 (97.3) |
|  | Posterior crosslink T6 | |  |
|  |  | False | 1444 (97.8) |
|  | Posterior crosslink T7 | |  |
|  |  | False | 1444 (97.8) |
|  | Posterior crosslink T8 | |  |
|  |  | False | 1450 (98.2) |
|  | Posterior crosslink T9 | |  |
|  |  | False | 1450 (98.2) |
|  | Posterior crosslink T10 | |  |
|  |  | False | 1463 (99.1) |
|  | Posterior crosslink T11 | |  |
|  |  | False | 1466 (99.3) |
|  | Posterior crosslink T12 | |  |
|  |  | False | 1446 (97.9) |
|  | Posterior crosslink L1 | |  |
|  |  | False | 1390 (94.1) |
|  | Posterior crosslink L2 | |  |
|  |  | False | 1318 (89.2) |
|  | Posterior crosslink L3 | |  |
|  |  | False | 1264 (85.6) |
|  | Posterior crosslink L4 | |  |
|  |  | False | 1178 (79.8) |
|  | Posterior fixation left T2 | |  |
|  |  | Uniplanar screw | 14 (0.9) |
|  |  | Polyaxial screw | 73 (4.9) |
|  |  | Screw | 24 (1.6) |
|  |  | Hook | 90 (6.1) |
|  | Posterior fixation left T3 | |  |
|  |  | Uniplanar screw | 106 (7.2) |
|  |  | Polyaxial screw | 180 (12.2) |
|  |  | Screw | 43 (2.9) |
|  |  | Hook | 269 (18.2) |
|  | Posterior fixation left T4 | |  |
|  |  | Uniplanar screw | 356 (24.1) |
|  |  | Polyaxial screw | 432 (29.2) |
|  |  | Screw | 76 (5.1) |
|  |  | Hook | 210 (14.2) |
|  | Posterior fixation left T5 | |  |
|  |  | Uniplanar screw | 783 (53.0) |
|  |  | Polyaxial screw | 268 (18.1) |
|  |  | Screw | 88 (6.0) |
|  |  | None | 74 (5.0) |
|  | Posterior fixation left T6 | |  |
|  |  | Uniplanar screw | 827 (56.0) |
|  |  | Polyaxial screw | 181 (12.3) |
|  |  | Screw | 95 (6.4) |
|  |  | None | 128 (8.7) |
|  | Posterior fixation left T7 | |  |
|  |  | Uniplanar screw | 943 (63.8) |
|  |  | Polyaxial screw | 123 (8.3) |
|  |  | Screw | 89 (6.0) |
|  |  | None | 87 (5.9) |
|  | Posterior fixation left T8 | |  |
|  |  | Uniplanar screw | 963 (65.2) |
|  |  | Polyaxial screw | 120 (8.1) |
|  |  | Screw | 90 (6.1) |
|  |  | None | 101 (6.8) |
|  | Posterior fixation left T9 | |  |
|  |  | Uniplanar screw | 1075 (72.8) |
|  |  | Polyaxial screw | 146 (9.9) |
|  |  | Screw | 93 (6.3) |
|  | Posterior fixation left T10 | |  |
|  |  | Uniplanar screw | 1152 (78.0) |
|  |  | Polyaxial screw | 144 (9.7) |
|  |  | Screw | 106 (7.2) |
|  | Posterior fixation left T11 | |  |
|  |  | Uniplanar screw | 1281 (86.7) |
|  |  | Polyaxial screw | 167 (11.3) |
|  | Posterior fixation left T12 | |  |
|  |  | Uniplanar screw | 1104 (74.7) |
|  |  | Polyaxial screw | 169 (11.4) |
|  |  | Screw | 119 (8.1) |
|  | Posterior fixation left L1 | |  |
|  |  | Uniplanar screw | 886 (60.0) |
|  |  | Polyaxial screw | 142 (9.6) |
|  |  | Screw | 88 (6.0) |
|  | Posterior fixation left L2 | |  |
|  |  | Uniplanar screw | 613 (41.5) |
|  |  | Polyaxial screw | 87 (5.9) |
|  |  | Screw | 65 (4.4) |
|  | Posterior fixation left L3 | |  |
|  |  | Uniplanar screw | 416 (28.2) |
|  |  | Polyaxial screw | 79 (5.3) |
|  |  | Screw | 44 (3.0) |
|  | Posterior fixation right T2 | |  |
|  |  | Uniplanar screw | 13 (0.9) |
|  |  | Polyaxial screw | 74 (5.0) |
|  |  | Screw | 19 (1.3) |
|  |  | Hook | 93 (6.3) |
|  | Posterior fixation right T3 | |  |
|  |  | Uniplanar screw | 68 (4.6) |
|  |  | Polyaxial screw | 183 (12.4) |
|  |  | Screw | 34 (2.3) |
|  |  | Hook | 262 (17.7) |
|  |  | None | 45 (3.0) |
|  | Posterior fixation right T4 | |  |
|  |  | Uniplanar screw | 254 (17.2) |
|  |  | Polyaxial screw | 367 (24.8) |
|  |  | Screw | 68 (4.6) |
|  |  | Hook | 234 (15.8) |
|  |  | None | 139 (9.4) |
|  | Posterior fixation right T5 | |  |
|  |  | Uniplanar screw | 589 (39.9) |
|  |  | Polyaxial screw | 254 (17.2) |
|  |  | Screw | 66 (4.5) |
|  |  | Hook | 72 (4.9) |
|  |  | None | 221 (15.0) |
|  | Posterior fixation right T6 | |  |
|  |  | Uniplanar screw | 754 (51.0) |
|  |  | Polyaxial screw | 156 (10.6) |
|  |  | None | 300 (20.3) |
|  | Posterior fixation right T7 | |  |
|  |  | Uniplanar screw | 840 (56.9) |
|  |  | Polyaxial screw | 123 (8.3) |
|  |  | Screw | 79 (5.3) |
|  |  | None | 179 (12.1) |
|  | Posterior fixation right T8 | |  |
|  |  | Uniplanar screw | 1010 (68.4) |
|  |  | Polyaxial screw | 114 (7.7) |
|  |  | None | 124 (8.4) |
|  | Posterior fixation right T9 | |  |
|  |  | Uniplanar screw | 1047 (70.9) |
|  |  | Polyaxial screw | 118 (8.0) |
|  |  | None | 130 (8.8) |
|  | Posterior fixation right T10 | |  |
|  |  | Uniplanar screw | 1040 (70.4) |
|  |  | Polyaxial screw | 134 (9.1) |
|  |  | None | 203 (13.7) |
|  | Posterior fixation right T11 | |  |
|  |  | Uniplanar screw | 1038 (70.3) |
|  |  | Polyaxial screw | 159 (10.8) |
|  |  | None | 242 (16.4) |
|  | Posterior fixation right T12 | |  |
|  |  | Uniplanar screw | 948 (64.2) |
|  |  | Polyaxial screw | 174 (11.8) |
|  |  | Screw | 109 (7.4) |
|  |  | None | 155 (10.5) |
|  | Posterior fixation right L1 | |  |
|  |  | Uniplanar screw | 865 (58.6) |
|  |  | Polyaxial screw | 163 (11.0) |
|  |  | Screw | 85 (5.8) |
|  | Posterior fixation right L2 | |  |
|  |  | Uniplanar screw | 597 (40.4) |
|  |  | Polyaxial screw | 105 (7.1) |
|  |  | Screw | 62 (4.2) |
|  | Posterior fixation right L3 | |  |
|  |  | Uniplanar screw | 410 (27.8) |
|  |  | Polyaxial screw | 87 (5.9) |
|  |  | Screw | 45 (3.0) |
|  | In-situ bending | |  |
|  |  | Yes | 325 (22.0) |
|  |  | No | 483 (32.7) |
|  | Rod tracing performed if no in-situ bending | |  |
|  |  | Yes | 215 (14.6) |
|  |  | No | 310 (21.0) |
|  |  | Null | 35 (2.4) |

Continuous predictors are summarized with mean and standard deviation (SD) and categorical predictors with total counts (N) and percentages (%).
